# Are Endoscopic Sinus Surgery Complication Rates Improving Over Time and With the Adoption of Image Guidance?

**DOI:** 10.1101/2025.11.07.25339508

**Authors:** Katherine Lauritsen, Arman Saeedi, Vijay R. Ramakrishnan

## Abstract

**Background:** Endoscopic sinus surgery (ESS) continues to be widely used as a treatment for refractory chronic rhinosinusitis (CRS). With advancements in technology and surgical technique, we might expect fewer surgical complications to occur in the modern era as compared to established rates that are often quoted in the literature and patient counseling.

**Methods:** We queried the TriNetX Research Network (2010-2023) for ESS complications (cerebrospinal fluid (CSF) leak, orbital injury, epistaxis event, ER visit, stroke, MI, thromboembolic event, 30-day mortality) by ICD and CPT codes in timeframes consistent with prior literature. Complication rates and IGS trends were evaluated over time (2010-2016 vs 2017-2023) and age groups (pediatric <12, adolescent (12-17), and adult >18-<65).

**Results:** Overall rate of major complications of CSF leak and orbital injury was similar or slightly better than prior reports, but epistaxis and other perioperative complications were higher than anticipated (>1%) and might deserve additional attention in preoperative counseling and perioperative care. IGS use increased over time and was associated with lower CSF leak rates (0.8 vs. 0.95%, p = 0.003), increased epistaxis rates (2.5 vs. 2.1%, p < 0.0001), and no significant difference in orbital injury. CSF leak was higher in patients under 18 (p <0.001), while adults had higher rates of epistaxis (p = 0.007).

**Conclusion:** Complications from ESS remain low. But increasing epistaxis and other perioperative complications are notable and may result from worsened overall population health. IGS use has increased with time and is associated with a decreased risk of CSF leak.

**Level of Evidence:** Level 4

**IRB:** IRB Protocol #27455 was deemed exempt from necessary IRB approval by the Indiana University School of Medicine Human Subjects Research Protection Program Board.

## Introduction

Endoscopic sinus surgery (ESS) is an integral part of modern otolaryngology practice. It is estimated that over a quarter of a million procedures are performed in the United States each year, most commonly as elective outpatient quality-of-life interventions for refractory chronic rhinosinusitis (CRS), but other indications do exist.^1^ Functional ESS was introduced in the U.S. in 1985, with known potential for major complications that appeared to decrease with surgeon experience.^2,3^ Since then, ESS has been widely taught and adopted as a standard of care in the management of medically refractory CRS.^4,5^

A foundational study of ESS complications reported major complications to occur in up to 8% of cases in an early cohort of 90 patients undergoing transnasal ethmoidectomies by a single surgeon in 1987.^3^ These major complications included hemorrhage, cerebrospinal fluid (CSF) leak, and orbital injury leading to blindness; minor complications included orbital injuries such as hematomas and orbital emphysema, adhesions, and ostial closure or stenosis. ^3^ With increasing individual and global experience in surgical technique, endoscopic anatomy, and technological advancements, complication rates inevitably declined. In 2012, a study using a national claims database reported an overall complication rate of 1.00%, with cerebrospinal fluid (CSF) leaks occurring in 0.17%, orbital injury in 0.07%, and hemorrhagic complications in 0.76% of 62,823 patients.^6^ These figures have become widely referenced in clinical discussions with patients regarding ESS risks despite these data being accumulated nearly 20 years ago.

As ESS continues to evolve, an updated query on complications may help our field understand if we have improved as a whole and identify any new trends or knowledge gaps to address. In addition to surgical experience, a specific enabling technology is the widespread availability of intraoperative computer-aided image guidance. Image-guided surgery (IGS) is used by Otolaryngologists to provide real-time feedback on the position of surgical instrumentation in relation to critical anatomical structures.^7^ The integration of IGS enhances education, anatomic understanding, surgical planning, completeness of surgery, and intraoperative confidence, but its benefits on surgical complication rates has been difficult to determine.^8,1^ Moreover, with the widespread adoption of image guidance in endoscopic sinus surgery, it remains important to discover if its use has resulted in fewer complications, an outcome that remains unclear as additional attention is now given to completeness of surgery and increasing case complexity among an aging and sicker population^9,10,1,11^

In recent years, several large databases have become accessible following large data collection efforts within electronic health records, which offer a new approach to examining these questions using appropriately powered cohorts. The objective of this study is to use a large nationally representative database of electronic medical record data (TriNetX Research Network) to examine rates of major complications following ESS, using real-world data to ensure that clinicians can adequately counsel patients on potential risks in the modern ESS era. Furthermore, we aim to assess whether the use of image guidance and patient age have any impact on the likelihood of experiencing these complications.

## Methods

### Study Design

This study was deemed exempt as “not human subjects research” by the Indiana University Institutional Review Board (IRB#27455). This retrospective cohort study utilized the TriNetX Research Network database, a large aggregate of de-identified electronic medical records from a variety of practice settings. The database contains information on patient demographics, diagnostic, and procedural data, updated in real-time. The database was queried for patients who underwent ESS between the years 2010-2023.

Consistent with prior research in this area, complications were identified using the International Classification of Diseases, 10^th^ edition (ICD-10) and Current Procedural Terminology (CPT) codes in the overall population undergoing surgery from 2010-2023. ^6,12–14^ Major postoperative complications were defined according to established categories of CSF leak, orbital injury, and hemorrhage requiring transfusion. “Other” perioperative complications, which are not universally referenced in existing literature on complications in sinus surgery, were included into a combined single measure, which included rates of unplanned emergency room visit, stroke, myocardial infarction, embolic or thrombosis event, and death within 30 days of ESS. Trends in the use of image guidance in ESS were also obtained, and its relationship to major complications was examined.

To mitigate confounding variables that may affect outcomes, propensity score matching was implemented as a function available within the TriNetX data analysis interface. Propensity matching was performed, including individual features such as age, BMI, and sex, and comorbid conditions such as diabetes, anticoagulation use, hypertension, nasal polyp status, and smoking status. While nasal polyp and smoking status were included in propensity matching, specific prevalence values for these variables were not extracted for reporting outside the matched analysis and, therefore, are not reported separately.

Prior reports have noted that complications may be different in pediatric patients.^15^ The cohort was stratified into three age groups: pediatric (< 12 years), adolescent (12-17 years), and adult (18-64 years), to evaluate if pediatric complication rates are different than adult ESS as has been suggested in prior literature. Major complication rates were also examined across two distinct time periods: 2010-2016 (Period 1) and 2017-2023 (Period 2), to assess potential temporal trends in complication rates and understand the adoption of IGS.

### Inclusion and Exclusion Criteria

The following CPT codes were used to identify individuals who underwent at least one of the following ESS procedures from 2010-2023: Ethmoidectomy: total with frontal sinus exploration: 31253, partial: 31254, total: 31255, including sphenoidotomy: 31257, with removal of tissue: 31259. Maxillary antrostomy: 31256, with removal of tissue: 31267. Frontal Sinus Exploration: 31276. Sphenoidotomy: 31287, with removal of tissue: 31288. Dilation of maxillary ostium: 31295, frontal sinus ostium: 31296, sphenoid sinus ostium: 31297, with frontal 31298. Endoscopy with biopsy, polypectomy, or debridement: 31237, with concha bullosa resection: 31240. The use of IGS: 61782 on the same day of surgery was also recorded.

The identification of major complications was completed by searching the ESS patient cohort for CPT codes and ICD-10 codes associated with the outcome. For CSF Leak: ICD-10 for CSF leak, postoperative leak, and meningitis: G96.0, G00, G03. CPT codes for placement of a lumbar drain: 62272 within 12 months of their first ESS surgery. For Orbital Injury: ICD-10 for unspecified visual disturbance, retrobulbar hematoma, orbital hemorrhage, or blindness: H53.19, H05.2, H54.X or CPT codes for canthotomy, canthoplasty, orbital decompression, or strabismus surgery-67900, 67901, 67500, 67311, 67312, 67314, 67316, within 6 weeks of ESS. For Epistaxis requiring transfusion: ICD-10 for epistaxis: R04.0 with CPT code for transfusion: 36430 within 3 months of first ESS. Although most complications were recognized and documented within the first few days after surgery, these postoperative time periods were chosen based on time-to-event analysis from our prior claims database study.^6^ Patients were excluded if they had documentation of any of these CPT or ICD-10 codes 12 months before their ESS date or if they were not included in the database for the 12 months following their first ESS surgery.

### Statistical Analysis

Statistical analysis was performed using the TriNetX integrated data analysis features, IBM SPSS Statistics (Version 29.0.2.0), and GraphPad PRISM (Version 10.4.2). Survival analysis was conducted using the Kaplan-Meier method to estimate event probability at the end of the designated time frame. The effect of IGS on event probability was assessed using the log-rank test. To estimate the relative risk of the event, a Cox proportional hazards regression model was used, and hazard ratios with 95% confidence intervals (CI) were reported. Chi-squared test for independence, as well as a pairwise z-test for proportions, was performed to assess the relationship between age groups and major complication rates, and the relationship between time period and major complication rates. A p-value of <0.05 was considered statistically significant.

## Results

A total of 150,775 patients who underwent ESS met the inclusion criteria. A summary of patient demographics and cohort characteristics is shown in **Table 1**. Intracranial complication was identified in 0.9% of cases, orbital injury in 0.2%, and epistaxis requiring transfusion in 2.3%. The rate of “other” complications, including unanticipated emergency room visit, stroke, heart attack, embolic or thrombosis event, and death within 30 days of ESS was 3.2% IGS was used in ∼43% of endoscopic sinus surgery cases over the 13-year period. In the overall cohort, there is a statistically significant lower risk of CSF leak with the use of IGS (.8% vs. .95%, p = 0.003), and there is no significant association between IGS use and the risk of orbital injury (**Table 2**). Conversely, IGS use was associated with an increased risk of significant epistaxis (2.5% vs. 2.1%, p <0.0001).

**Table 1:** Patient Demographics and Cohort Characteristics.

|  | Overall Cohort<br>N= 150,775 | Adult Cohort (>18-<65<br>Years) N=143,542 | Adolescent Cohort<br>(>12-<18 Years)<br>N=6386 | Pediatric Cohort<br>(<12 Years)<br>N=5435 |
| --- | --- | --- | --- | --- |
| Mean Age (± SD) | 47.8 (19.2) | 50.3 (16.3) | 14.7 (1.71) | 6.44 (3.25) |
| Sex (N, % Male) | 74340 (49) | 69671 (49) | 3698 (58) | 3237 (60) |
| BMI | 28.6 | 29 | 23.9 | 18.1 |
| Race (% Total) |  |  |  |  |
| - White | - 67 | - 68 | - 64 | - 66 |
| - Hispanic | - 8 | - 7 | - 18 | - 16 |
| - Black | - 10 | - 9 | - 12 | - 12 |
| - Asian | - 4 | - 4 | - 2 | - 2 |
| - Other | - 0 | - 0 | - 0 | - 0 |
| Type 2 Diabetics<br>(N, % Total) | 15515 (10) | 16155 (11) | 84 (1) | 23 (0) |
| Hypertensive<br>Diseases (N, %<br>Total) | 40714 (27) | 41879 (29) | 232 (4) | 239 (4) |
| Anticoagulation or<br>Platelet Aggregate<br>Inhibitors Use (N,<br>% Total) | 52939 (35) | 52649 (37) | 683 (11) | 622 (12) |

**Table 2:** Overall complication rates, with and without the use of IGS from 2010-2023 in matched cohort.

|  | CSF Leak (% Risk) | Orbital Injury (% Risk) | Hemorrhage (% Risk) |
| --- | --- | --- | --- |
| Total | 1372/144638 (.9%) | 331/ 145694 (.23%) | 3180 /140254 (2.3%) |
| No IGS | 592/62036 (.95%) | 118/ 63590(.19%) | 1223/61677 (2.1%) |
| IGS | 515/63049 (.8%) | 140/62542 (.24%) | 1539/59566 (2.5%) |
| Hazard Ratio (Confidence Interval) | **0.8 (0.746,0.945)** | 1.2 (0.926, 1.51) | **1.2 (1.11, 1.30)** |
| Percentage Total IGS Used | 43.6% | 42.9% | 42.7% |

CSF= Cerebrospinal Fluid; IGS=Image-guided Surgery.

The adoption of IGS increased from 37.9% of all surgical cases in Period 1 (2010-2016) to 43.8% in Period 2 (2017-2023) (p<0.001, **Table 3**). While the rate of intracranial complications remained relatively stable across the two periods (1.0 vs. 0.9%, p= 0.8), orbital injury decreased significantly (0.3% in Period 1 vs 0.2% in Period 2, p<.001). Conversely, the rate of hemorrhage increased in the more recent period (2.0 vs 2.4 %, p <.001) (**Table 4**).

**Table 3.** Number of endoscopic sinonasal surgeries overall that used IGS in different time-periods.

| Time Periods | 2010-2016 | 2017-2023 |
| --- | --- | --- |
| IGS Used | 20,171 | 46,976 |
| IGS Not Used | 33,103 | 60,390 |
| **P= <.001** |  |  |

IGS= Image-guided surgery

**Table 4:** Overall Rates of Complications Across Two Time Periods in a matched cohort.

|  | CSF Leak (%Risk) | Orbital Injury (% Risk) | Hemorrhage (% Risk) |
| --- | --- | --- | --- |
| 2010-2016 | 513/51165 (1.00%) | 144/50838 (.28%) | 992/49866 (1.99%) |
| 2017-2023 | 936/102780 (.91%) | 206/105219 (.2%) | 2381/99133 (2.4%) |
|  | p= .0784 | **p= 0.0006** | **p= <.0001** |

CSF= Cerebrospinal Fluid

When stratifying the data by age group, the rate of complications varied significantly across the groups. Specifically, risk of CSF leak was higher in those under 18 years old, with a 0.3-0.7% increased risk (p <0.001). There was no statistically significant difference in the rate of orbital injury in regard to age (p= 0.04, CI -0.05%-0.16%, Z= 0.8) (**Table 5**). Adult patients (18-64 years) had a significantly higher risk of hemorrhage compared to younger patients (0.1-0.6%; p= 0.007).

**Table 5:** Overall complication rates by age group from 2010-2023 in matched cohort.

|  | CSF Leak (% Risk) | Orbital Injury (% Risk) | Hemorrhage (% Risk) |
| --- | --- | --- | --- |
| <12 years | 85/5149 (1.7%) | 20/5209 (.38%) | 98/4898 (2.0%) |
| 12-18 years | 77/6064 (1.3%) | <10/6084 (.164%) | 104/5770 (1.8%) |
| >18 - <65 years | 1238/136741 (.9%) | 305/138766 (.22%) | 3047/132956 (2.3%) |
|  | **p <0.001** | **p= .021** | **p=.023** |

CSF= Cerebrospinal Fluid

## Discussion

Because ESS complications occur infrequently, we used a large national database study design to achieve the requisite statistical power for our inquiries, while also broadly representing surgeons, patients, and operative environments in the real world.^1,16^ This study, with over 150,000 ESS patients spanning thirteen years, reveals several novel and clinically meaningful trends. First, we provide more recent and new information on major preoperative complication rates that can be used for shared decision making and preoperative counseling. Intracranial and orbital complications remain rare, and perhaps are slightly improving over time. A second key finding is that, in contrast to intracranial and orbital complication rates, significant postoperative epistaxis was higher than expected (2.3%), exceeding rates in prior reports. ^2,6,12^ This may be due to expanding indications for sinus surgery in at-risk populations (e.g., immunosuppressed, ICU populations)^17^, extent of surgery, or increased anticoagulation. Indeed, we serve an aging patient population with more comorbid conditions,^18,19^ including a markedly increased anticoagulation use. Similar trends have been reported in dental and GI surgical procedures, where patients resuming anticoagulant therapy had an increase in bleeding risk despite medications being held preoperatively. ^20,21^ Further work is necessary to illuminate whether this observation reflects this U.S. healthcare trend, is a function of changes in operative strategy, or is inherent to varied coding and reimbursement approaches whereby surgeons may use FESS codes for extended cases (rather than unlisted 31299 codes^22^).

Second, we found that the use of image guidance in FESS was associated with a decreased risk of CSF leaks but a minor increased risk of postoperative epistaxis. Moreover, its use continued to grow over time, suggesting that surgeons believe it confers significant advantages and/or there is a trend towards more extensive or complex surgeries being performed where the use of IGS is beneficial.^13^ The influence of IGS on surgical outcomes has been difficult to demonstrate, for a number of reasons that are described in the literature.^1,8,11,16^ Krings et al. reported that IGS was associated with a higher likelihood of major complications, examining state payer databases from 20025-2008 (Odds ratio [OR] 1.95, 95% confidence interval [CI] 1.42-2.66).^12^ Since then, other studies have shown no association or a slight positive effect for IGS and ESS complications. Dalgorf et al., ^23^ performed a systematic review and meta-analysis of IGS in ESS and found that major complications and total complications are less likely to occur with the use of IGS. The authors argue that allocation bias of more difficult surgeries to include IGS, further supports its possible benefit on ESS complication rates. Although the effect size in our results is modest, the statistically significant finding supports reimbursement policies that recognize the added benefit of IGS in FESS cases, when indicated. ^24,25^

Third, whereas prior database studies conflicted in regards to major complication rates by age, we observed that CSF leak and orbital injury occurred more frequently in patients under 18. This may be related to acute infectious complications of sinusitis are a common indication for ESS in pediatric patients, challenges of pediatric anatomy, or paranasal sinus pneumatization patterns in children. Notably, greater than 10% of the pediatric cohort was on anticoagulation therapy, indicating the degree of medical complexity in pediatric patients undergoing ESS. Krings et al. reported higher complication rates with increasing age, but in Ramakrishnan et al., it was observed that orbital complications may occur more frequently in young children ^6^.^12^

Taken together, these data indicate that ESS remains safe in terms of major complications, but that postoperative epistaxis and perioperative medical complications may be more common than previously recognized. The latter is of particular relevance given population-level trends with increasing age, comorbid conditions, use of anticoagulation, and the skull/orbit plateau. Updated estimates for complication rates of “all-comers” are meaningful foundations for discussion in shared decision-making for surgery, especially when the indication is for elective quality-of-life improvement. Prior research has shown that the majority of patients considering ESS are interested to learn about complications that occur at a frequency >1%.^26^ We suggest that preoperative discussions include updated numbers for risk of postoperative bleeding and “other” unexpected events, such as unplanned emergency room visit or medical event, particularly when comorbid health conditions are present.

This study’s strengths include the use of the large, contemporary TriNetX medical records database, which has received increasing interest because of its large cohort size, broad representation, up-to-date information, open access, and intuitive user interface. However, one of its inherent limitations is the inability to access raw patient data for more granular details and advanced statistical analyses to account for surgery indication, extent of surgery, anatomic complexity, disease severity, or surgeon experience. Individual surgeons may have different approaches to coding, operative technique, use of particular instrumentation and biomaterials, and follow-up. Although it is of interest to evaluate complication rates by operative indication and extent of surgery, the numerous permutations would inevitably result in loss of statistical power. ^1,16^ We applied a strict and conservative approach to capturing postoperative events by coding and time period, one that was informed by time-to-event analyses from our prior study.^6^ However, it is possible that some events captured were not a direct result of surgery.

Future research may explore some of these considerations, including the varied indications for ESS, newer measures of disease severity and extent of surgery, and the application of technologies such as IGS, hemostatic biomaterials, or balloons. Additionally, the impact of healthcare resource availability, geographic setting, or socioeconomic status is of interest as new metrics and databases become available. Lastly, it may be valuable to consider if it is possible to continue to improve our ESS complication rates or if we have reached a plateau, and where our future focus should lie.

## Conclusion

Major complication rates arising from ESS remain rare, but significant postoperative hemorrhage appears to be increasing in tandem with an aging and anticoagulated patient population. However, stable CSF leak and orbital injury rates suggest that surgical technique and training are adapting to the increasing patient complexity and expanding surgical indications. IGS use is likely used in more complex surgical cases, and is associated with fewer intracranial complications, although a statistical increase in postoperative hemorrhage. These findings provide data to guide shared decision-making, inform reimbursement discussions, and direct research and value-based discussions on understanding potential strategies to reduce surgical complications.

## Data Availability

All data produced in this manuscprit is available to researchers with subscribed access to the TriNetX Research Network.

## Acknowledgments

Guidance on biostatistical analysis and data interpretation was provided by Dr. Erik Parker, PhD, IU Bloomington School of Public Health.

